# Risk factors, outcomes, and predictors of therapeutic response in preterm infants with patent ductus arteriosus: A retrospective cohort study

**DOI:** 10.64898/2026.04.10.26350668

**Authors:** Hayet Ben Hamida, Maroua El Ouaer, Sioir Abdelmoula, Maha El Ghali, Manel Bizid, Ikram Chamtouri, Kamel Monastiri

## Abstract

**Background:** Patent ductus arteriosus (PDA) is a common and potentially serious cardiovascular condition in preterm infants, particularly those with low gestational age and birth weight. Its management remains controversial due to variability in screening, diagnostic criteria, and treatment strategies. This study aimed to evaluate risk factors, outcomes, and management strategies for PDA in preterm infants, and to identify predictors of clinical and echocardiographic response to therapy.

**Methods:** We conducted a retrospective cohort study over a 4-year period (2016–2019) in the neonatal intensive care unit (NICU) of a tertiary care center. All consecutive preterm infants admitted during the study period were eligible. Infants with echocardiographically confirmed PDA who received pharmacological treatment with intravenous paracetamol or ibuprofen were included in the analysis. Missing data were minimal and handled using available-case analysis. Statistical analyses included descriptive statistics, Pearson’s chi-square test, and multivariable logistic regression.

**Results:** Among 2154 preterm infants admitted to the NICU, 60 were diagnosed with PDA (incidence : 2.8%). The mean gestational age was 29 ± 2.6 weeks, and the median birth weight was 1200 g. Respiratory distress occurred in 95% of cases, mainly due to hyaline membrane disease (86.7%). PDA was symptomatic in 80% of infants. First-line treatment resulted in clinical improvement in 77% and ductal closure in 83.3% of cases, most within 3 days. Predictors of successful closure included gestational age ≥ 28 weeks (OR = 5.9; 95% CI : 1.7–20.2) and antenatal corticosteroid exposure (OR = 1.2; 95% CI : 1.0–1.6). Overall mortality was 35% and was significantly higher in infants < 28 weeks (OR = 5.0; 95% CI : 2.4–10.3). Clinical improvement (OR = 3.7) and echocardiographic closure (OR = 4.5) after first-line treatment were associated with reduced mortality.

**Conclusions:** PDA in preterm infants is associated with substantial morbidity and mortality, particularly in those born before 28 weeks of gestation. Early diagnosis, antenatal corticosteroid exposure, and timely pharmacological treatment may improve outcomes. Systematic echocardiographic screening in high-risk neonates should be considered.

## Introduction

Patent ductus arteriosus (PDA) is one of the most common cardiovascular conditions in preterm neonates, with an incidence ranging from 30% to 60% [1]. It affects nearly one-third of infants born before 30 weeks of gestation and up to 70% of those born before 28 weeks [2]. Its incidence is inversely related to gestational age and birth weight [3].

Under physiological conditions, the ductus arteriosus undergoes functional closure within the first 72 hours of life, followed by anatomical closure. This process is primarily triggered by lung expansion and the cessation of placental circulation [4]. However, in preterm infants, this mechanism is often impaired, leading to persistent ductal patency.

Major risk factors for PDA include low gestational age, very low birth weight, neonatal respiratory distress—mainly due to hyaline membrane disease—surfactant administration, and perinatal infection [5]. Persistent ductal patency results in left-to-right shunting, causing pulmonary overcirculation and systemic hypoperfusion. The clinical consequences depend on the magnitude and hemodynamic significance of the shunt [6].

PDA is associated with several serious complications, including pulmonary hemorrhage [7], intraventricular hemorrhage, necrotizing enterocolitis, and renal hypoperfusion [5]. It therefore represents a significant contributor to neonatal morbidity and mortality. An Italian study reported a mortality rate of 10.4% in cases of hemodynamically significant PDA [8]. Moreover, PDA increases the risk of neurological sequelae, particularly in infants requiring pharmacological (1.6-fold increase) or surgical treatment (twofold increase) [9].

Despite its clinical importance, the management of PDA remains controversial, particularly in settings where systematic echocardiographic screening is not routinely implemented. While indomethacin was historically the standard treatment [10], ibuprofen and paracetamol are currently the most widely used pharmacological options.

In this context, the present study aimed to evaluate risk factors, morbidity, and mortality associated with PDA in preterm infants, and to identify predictors of response to pharmacological treatment.

## Methods

### Study design and setting

We conducted a retrospective, single-center cohort study over a 4-year period (January 2016 to December 2019) in the neonatal intensive care unit of a tertiary care center specializing in high-risk neonates. All consecutive preterm infants admitted during the study period were screened for eligibility.

### Study population

A total of 2154 preterm infants admitted between January 2016 and December 2019 were screened. Among these, infants with echocardiographically confirmed patent ductus arteriosus (PDA) who received pharmacological treatment with intravenous paracetamol or ibuprofen were included (n = 60).

Infants with congenital heart disease other than PDA, those managed conservatively without pharmacological treatment, and those without echocardiographic confirmation were excluded.

### Diagnosis and follow-up

The diagnosis of PDA was based on echocardiographic parameters, including ductal diameter, shunt direction, left atrium-to-aorta (LA/Ao) ratio, and Doppler flow patterns. Echocardiographic assessments were performed according to standardized protocols routinely applied in the unit and consistent with established pediatric cardiology guidelines.

Examinations were conducted by experienced operators (pediatric cardiologists and/or trained neonatologists). To minimize inter-observer variability, predefined measurement criteria were applied.

Hemodynamically significant PDA was defined by a ductal diameter ≥ 1.5 mm, an LA/Ao ratio ≥ 1.5, and/or clinical signs of systemic hypoperfusion.

Follow-up echocardiography was performed after the first course of treatment. In cases of persistent PDA, a second course using the same or an alternative drug was considered unless contraindicated.

### Treatment protocol

Ibuprofen was administered at a dose of 10 mg/kg on day 1, followed by 5 mg/kg on days 2 and Paracetamol was administered at a dose of 15 mg/kg every 6 hours for 3 to 5 days.

The choice of treatment was based on clinical condition, contraindications, and physician discretion.

### Data collection

Data were extracted from medical records and echocardiographic reports using a standardized data collection form

Collected variables included perinatal characteristics (gestational age, birth weight, sex, antenatal corticosteroid exposure, maternal conditions, and mode of delivery), neonatal clinical course (respiratory distress, surfactant use, mechanical ventilation, and hemodynamic status), PDA characteristics (ductal size, shunt direction, LA/Ao ratio, and pulmonary hypertension), treatment response (clinical improvement and echocardiographic closure), and outcomes (bronchopulmonary dysplasia, intraventricular hemorrhage, necrotizing enterocolitis, and survival to discharge).

### Definitions

Clinical improvement was defined as a reduction in respiratory distress, improved oxygenation, and/or hemodynamic stabilization following treatment. Treatment failure was defined as persistence of hemodynamically significant PDA after the first course of pharmacological treatment, confirmed by echocardiography.

Hypotrophy was defined as birth weight below the 10th percentile for gestational age. Very preterm birth was defined as gestational age < 28 weeks.

### Statistical analysis

Statistical analyses were performed using SPSS version 21. Continuous variables were expressed as mean ± standard deviation or median (interquartile range), as appropriate, and categorical variables as percentages.

Univariate analysis was performed using Pearson’s chi-square test to identify factors associated with outcomes. Variables with a p-value < 0.20 and those considered clinically relevant based on previous literature were included in multivariable logistic regression models. A stepwise selection procedure was applied to identify independent predictors in the final model, including gestational age, birth weight, antenatal corticosteroid exposure, and neonatal clinical status. A p-value < 0.05 was considered statistically significant.

### Sample size

All eligible cases during the study period were included. No formal sample size calculation was performed due to the retrospective design.

### Bias and confounding

Potential sources of bias were considered. Selection bias may have arisen from the single-center retrospective design and inclusion of only treated cases, potentially excluding milder PDA forms. This was mitigated by screening all consecutive preterm infants during the study period.

Information bias was minimized through standardized data extraction and predefined echocardiographic criteria. However, the involvement of multiple operators may have introduced some inter-observer variability.

Confounding was addressed using multivariable logistic regression including key clinical variables such as gestational age, birth weight, antenatal corticosteroid exposure, and neonatal clinical status.

### Ethics statement

This retrospective study was conducted using anonymized medical records of preterm infants hospitalized between January 2016 and December 2019. The requirement for informed consent was waived by the institutional ethics committee.

All procedures were performed in accordance with the principles of the Declaration of Helsinki and relevant national regulations. All data were fully anonymized prior to analysis, and no identifiable information was accessible to the investigators.

## Results

### Study population

During the study period, 2154 preterm infants were admitted to the neonatal intensive care unit (NICU) and screened for eligibility. Among them, 60 infants with echocardiographically confirmed patent ductus arteriosus (PDA) who received pharmacological treatment were included, corresponding to an incidence of 2.8% (60/2154). No further exclusions were applied after initial screening. Missing data were minimal and handled using available-case analysis.

Delivery was performed by emergency cesarean section in 56.7% (n = 34). The mean gestational age was 29 ± 2.6 weeks (range : 28–32+6 weeks), and the median birth weight was 1200 g (range : 620–3000 g). The male-to-female ratio was 1.14. Gestational toxemia was the most frequent maternal condition (53.3%, n = 32), including pre-eclampsia in 21.7% (n = 13). Antenatal corticosteroids were administered in 50% of pregnancies (n = 30).

### Neonatal clinical features

Neonatal respiratory distress (NRD) occurred in 95% of infants (n = 57), most frequently due to hyaline membrane disease (86.7%, n = 52). Early-onset neonatal infection was observed in 15% (n = 9), while pneumothorax and transient tachypnea of the newborn occurred in 10% (n = 6) and 5% (n = 3), respectively. Surfactant therapy was administered in 70% of neonates (n = 42). Mechanical ventilation was required in 59 infants, predominantly invasive (83.1%, n = 49), with non-invasive ventilation in 16.9% (n = 10). The distribution of underlying etiologies and treatment modalities for NRD is summarized in **Table 1**.

**Table 1.** Etiologies and management of neonatal respiratory distress syndrome.

| Category | Variable | Frequency (n) | Percentage (%) |
| --- | --- | --- | --- |
| **Etiologies** | Hyaline membrane disease | 52 | 86.7 |
|  | Early-onset neonatal infection | 9 | 15.0 |
|  | Pneumothorax | 6 | 10.0 |
|  | Transient tachypnea of the newborn | 3 | 5.0 |
| **Treatment** | Surfactant administration | 42 | 70.0 |
|  | Ventilatory support (n = 59) |  |  |
|  | Non-invasive ventilation | 10 | 16.9 |
|  | Invasive mechanical ventilation | 49 | 83.1 |

**Notes:** Percentages may exceed 100% due to multiple diagnoses per patient. Percentages for ventilatory support are calculated using n = 59 as the denominator.

Among infants with NRD receiving pharmacological treatment, 88% (n = 53) required a single course, while 12% (n = 7) required two courses.

PDA was asymptomatic in 11.7% (n = 7), mainly in the presence of risk factors such as extreme prematurity, severe NRD requiring surfactant, and absence of antenatal corticosteroids. In the remaining 88.3% (n = 53), PDA was diagnosed following clinical deterioration. Key respiratory and hemodynamic signs are presented in **Table 2**. Most frequent respiratory signs included increased oxygen requirements (73.3%, n = 44), apnea (35%, n = 21), and alveolar hemorrhage (13.3%, n = 8). Hemodynamic instability was common, with blood pressure abnormalities in 53.3% (n = 32), tachycardia in 41.7% (n = 25), volume expansion required in 46.7% (n = 28), and vasoactive drugs used in 31.7% (n = 19).

**Table 2.** Clinical signs suggestive of patent ductus arteriosus.

| Category | Clinical Sign | Frequency (n) | Percentage (%) |
| --- | --- | --- | --- |
| **Respiratory signs** | Apneas | 21 | 35.0 |
|  | Alveolar hemorrhage | 8 | 13.3 |
|  | Secondary increase in O<sub>2</sub> needs | 44 | 73.3 |
| **Hemodynamic signs** | Heart murmur | 13 | 21.7 |
|  | Blood pressure differential | 32 | 53.3 |
|  | Tachycardia | 25 | 41.7 |
|  | Vascular filling | 28 | 46.7 |
|  | Use of vasoactive drugs | 19 | 31.7 |

**Notes :** Signs can be combined

### Echocardiographic findings

Echocardiography was performed within the first 24 hours in 26.7% of neonates (n = 16), on day 2 in 35% (n = 21), on day 3 in 20% (n = 12), and after day 3 in 18.3% (n = 11). The mean PDA diameter was 3.2 ± 0.84 mm (range : 1.9–5 mm), with a diameter ≥ 1.5 mm in 96.7% (n = 58). The LA/Ao ratio was measured in 30% (n = 18), with a mean of 1.69 ± 0.43.

The ductal shunt was left-to-right in 75% (n = 45), bidirectional in 20% (n = 12), and right-to-left in 3.3% (n = 2) (**Figures 1 and 2**). Additional echocardiographic details, including PDA diameter intervals, LA/Ao ratio, ductal shunt, and pulmonary hypertension, are summarized in **Table 3**. Pulmonary hypertension was present in 18.3% of neonates (n = 11).

**Figure 1.**
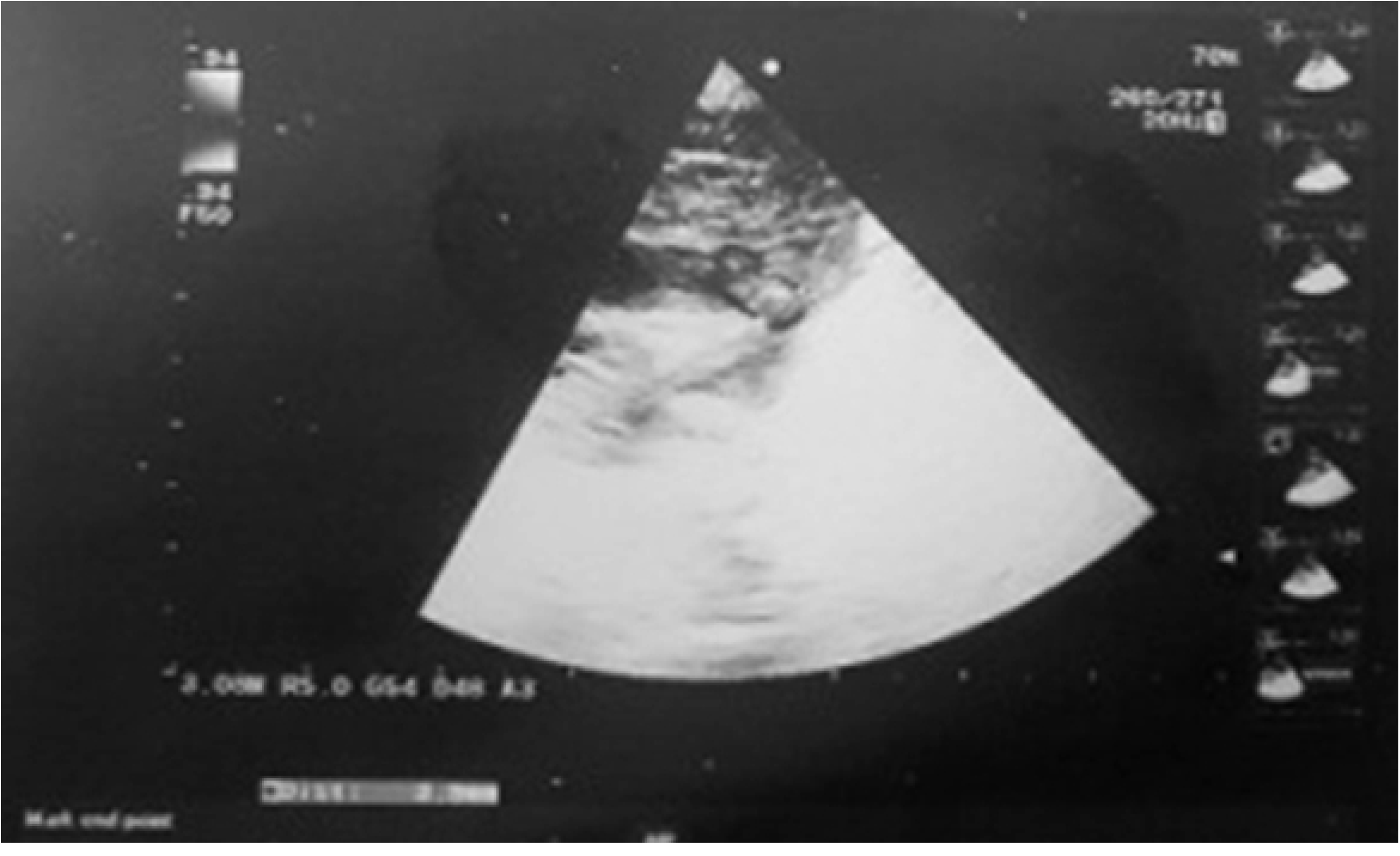
Small axis parasternal section showing a broad ductus arteriosus with color Doppler specifying the nature of the left to right shunt.

**Figure 2.**
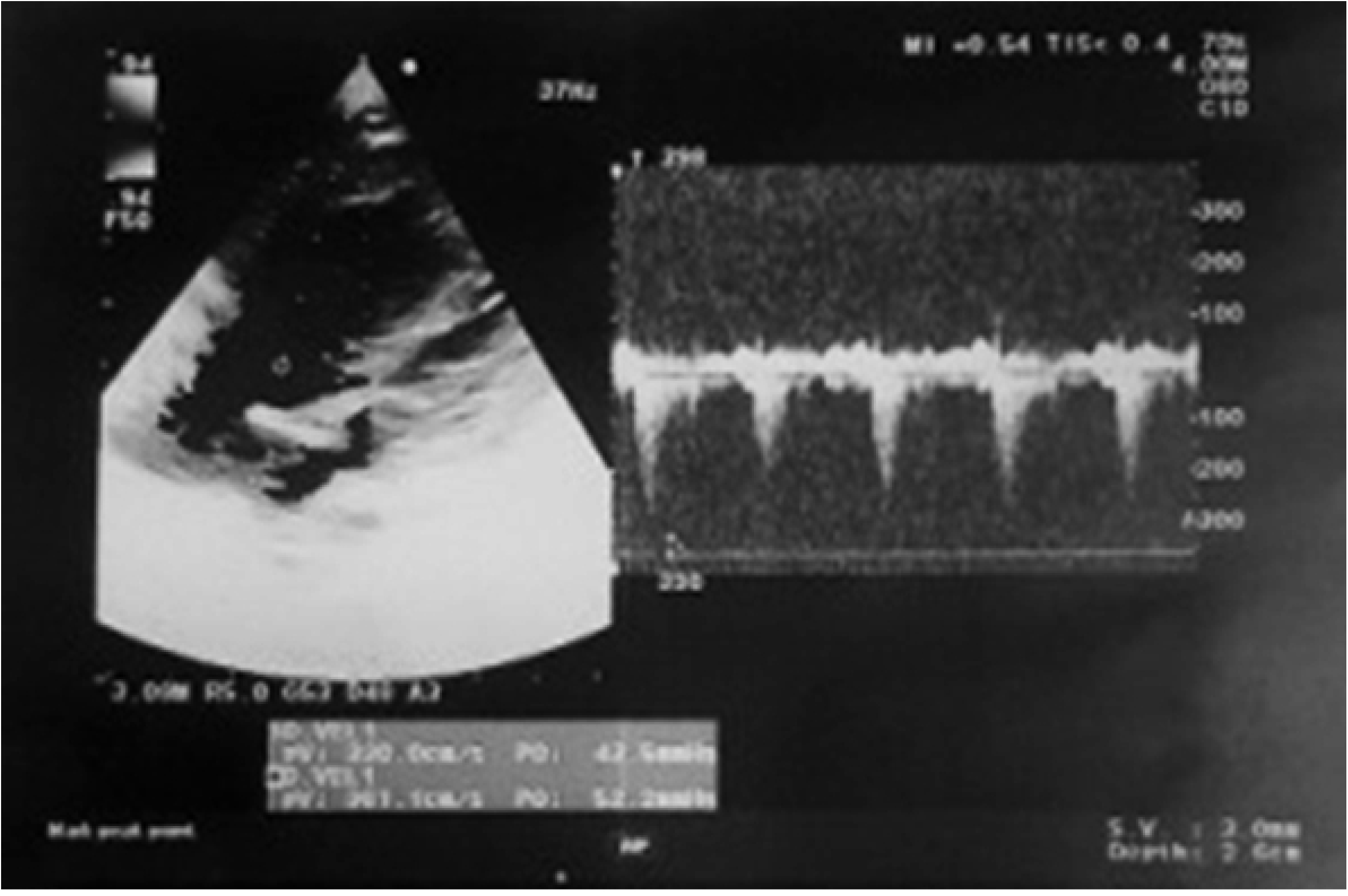
Pulsed Doppler specifying velocity through the ductus arteriosus.

**Table 3.**
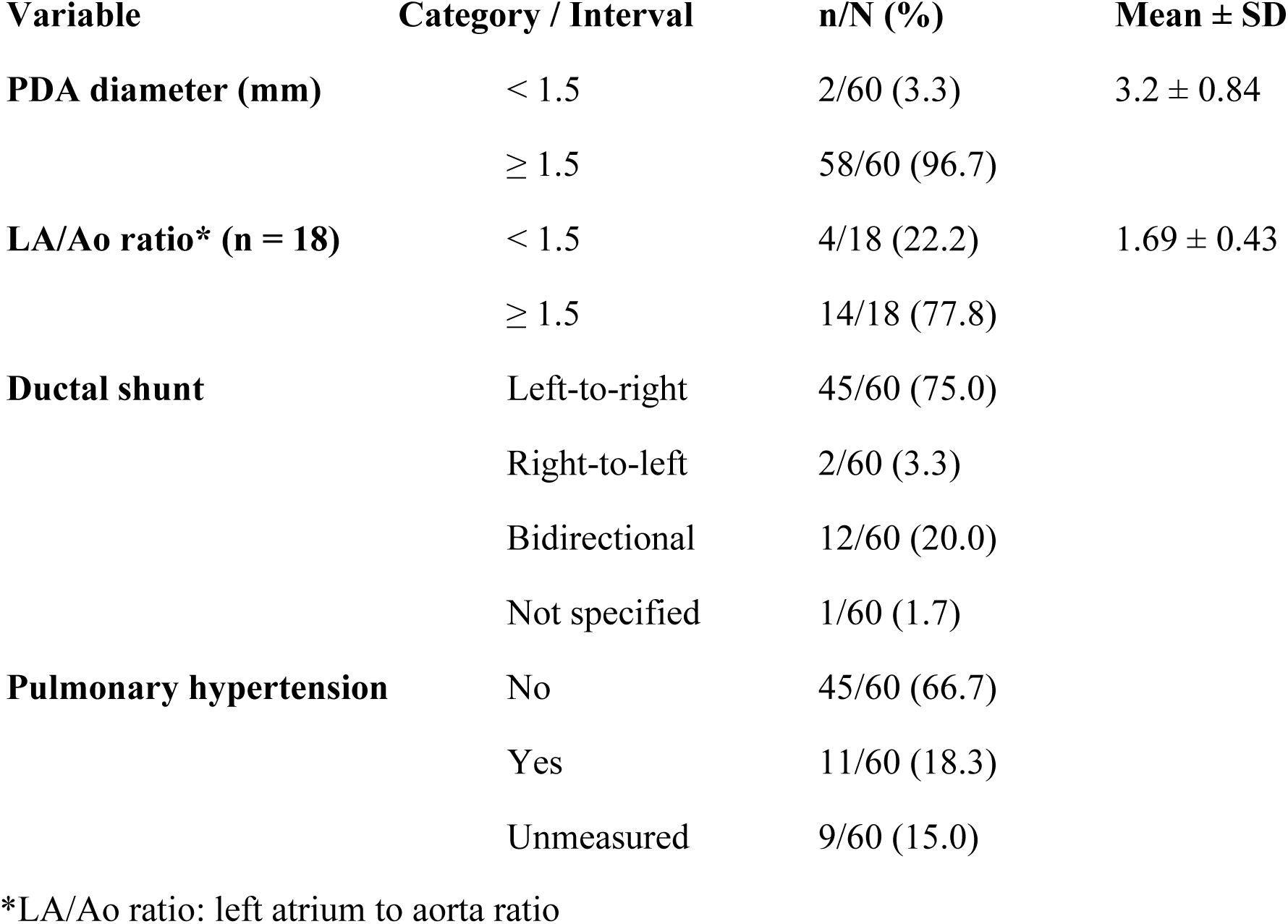
Echocardiographic characteristics of neonates with patent ductus arteriosus.

| **Variable** | **Category / Interval** | **n/N (%)** | **Mean ± SD** |
| --- | --- | --- | --- |
| **PDA diameter (mm)** | < 1.5 | 2/60 (3.3) | 3.2 ± 0.84 |
|  | ≥ 1.5 | 58/60 (96.7) |  |
| **LA/Ao ratio\* (n = 18)** | < 1.5 | 4/18 (22.2) | 1.69 ± 0.43 |
|  | ≥ 1.5 | 14/18 (77.8) |  |
| **Ductal shunt** | Left-to-right | 45/60 (75.0) |  |
|  | Right-to-left | 2/60 (3.3) |  |
|  | Bidirectional | 12/60 (20.0) |  |
|  | Not specified | 1/60 (1.7) |  |
| **Pulmonary hypertension** | No | 45/60 (66.7) |  |
|  | Yes | 11/60 (18.3) |  |
|  | Unmeasured | 9/60 (15.0) |  |

\*LA/Ao ratio: left atrium to aorta ratio

### Treatment outcomes

Paracetamol was used in 73.3% (n = 44), while ibuprofen was used in 26.7% (n = 16). Clinical improvement after first-line treatment was observed in 77% (n = 46), and echocardiographic closure was achieved in 83.3% (n = 50), with 80% closing within 3 days and 20% within 5 days. A second treatment course was required in 10% (n = 6). No ductal reopening was observed. Factors associated with clinical response to first-line treatment are presented in **Table 4**, showing that treatment failure was significantly associated with hypotrophy (p = 0.028) and absence of antenatal corticosteroids (p = 0.011).

**Table 4.** Prognostic factors associated with clinical response to first-line treatment.

| **Variable** | **Category** | **No improvement**<br>**(n=7)** | **Improvement**<br>**(n=53)** | **p-**<br>**value** |
| --- | --- | --- | --- | --- |
| **Trophic status** | No hypotrophy | 3 (33.3) | 6 (66.7) | **0.028** |
|  | Hypotrophy | 4 (7.8) | 47 (92.2) |  |
| **Gestational age** | < 28 weeks | 4 (23.5) | 13 (76.5) | 0.092 |
|  | ≥ 28 weeks | 3 (7.0) | 40 (93.0) |  |
| **Lung maturation** | Not done | 7 (23.3) | 23 (76.7) | **0.011** |
|  | Done | 0 (0.0) | 30 (100.0) |  |
| **Surfactant therapy** | Not given | 2 (11.1) | 16 (88.9) | 0.999 |
|  | Given | 5 (11.9) | 37 (88.1) |  |
| **Early-onset neonatal infection (EONI)** | No | 4 (10.3) | 35 (89.7) | 0.687 |
|  | Yes | 3 (14.3) | 18 (85.7) |  |
| **DA diameter (mm)** | < 1.5 | 0 (0.0) | 2 (100.0) | 0.999 |
|  | ≥ 1.5 | 7 (12.1) | 51 (87.9) |  |
| **Hemodynamically significant DA** | No | 0 (0.0) | 10 (100.0) | 0.319 |
|  | Yes | 7 (17.9) | 32 (82.1) |  |
| **Treatment used** | Perfalgan® | 5 (11.4) | 39 (88.6) | 0.999 |
|  | PEDEA® | 2 (12.5) | 14 (87.5) |  |
| **Age at treatment initiation** | ≤ 24 hours | 1 (7.1) | 13 (92.9) | 0.999 |
|  | > 24 hours | 6 (13.0) | 40 (87.0) |  |

Multivariate logistic regression identified independent predictors of outcomes after adjustment for gestational age, birth weight, antenatal corticosteroid exposure, and neonatal clinical status. Antenatal corticosteroids increased the likelihood of clinical improvement (OR = 2.3; 95% CI : 1.6–3.1). PDA closure after the first treatment course was significantly associated with gestational age ≥ 28 weeks (OR = 5.9; 95% CI : 1.7–20.2 ; p = 0.003). Antenatal lung maturation showed a trend toward association with PDA closure (OR = 5.1; 95% CI : 0.96–26.9 ; p = 0.038) (**Table 5**).

**Table 5.** Multivariate analysis of factors associated with ductal closure after first-line treatment.

| Variable | Category | Closure failure (n = 10) | Closure (n = 50) | p-value | OR (95% CI) |
| --- | --- | --- | --- | --- | --- |
| Gestational age | < 28 weeks | 7 (41.2%) | 10 (58.8%) | 0.003 | 5.9 (1.7–20.2) |
|  | ≥ 28 weeks | 3 (7.0%) | 40 (93.0%) |  | Reference |
| Lung maturation | Not given | 8 (26.7%) | 22 (73.3%) | 0.038 | 5.1 (0.96–26.9) |
|  | Given | 2 (6.7%) | 28 (93.3%) |  | Reference |

**Abbreviations:** OR, odds ratio; CI, confidence interval.
**Reference category:** ≥28 weeks for gestational age; lung maturation given.

### Short-term complications and mortality

Bronchopulmonary dysplasia occurred in 5% (n = 3), with no significant associations. Intraventricular hemorrhage occurred in 36.7% (n = 22) and was significantly associated with gestational age < 28 weeks (p = 0.038). Necrotizing enterocolitis occurred in 11.7% (n = 7), showing borderline associations with hypotrophy (p = 0.062) and hemodynamically significant PDA (p = 0.051).

A total of 21 deaths (35%) were recorded, including 8 during the first week. Mortality was significantly higher in infants born before 28 weeks (82.4% vs. 16.3% ; p < 0.001). Independent predictors of mortality included gestational age < 28 weeks (OR = 5 ; 95% CI : 2.4–10.3), lack of clinical improvement after first-line treatment (OR = 3.7; 95% CI : 2.4–5.9 ; p < 0.001), and absence of echocardiographic closure after the first course (OR = 4.5; 95% CI : 2.6–7.6 ; p < 0.001) (**Table 6**).

**Table 6.** Multivariate analysis of factors associated with mortality related to patient management.

| Variable | Category | Survival (n = 39) | Death (n = 21) | p-value | OR (95% CI) |
| --- | --- | --- | --- | --- | --- |
| Clinical response to treatment (first-line) | No improvement | 0 (0%) | 7 (100%) | <0.001 | 3.7 (2.4–5.9) |
|  | Improvement | 39 (73.6%) | 14 (26.4%) |  | Reference |
| Echocardiographic closure of the ductus arteriosus (first course) | No | 0 (0%) | 10 (100%) | <0.001 | 4.5 (2.6–7.6) |
|  | Yes | 39 (78%) | 11 (22%) |  | Reference |

**Abbreviations:** OR, odds ratio; CI, confidence interval
**Reference category:** Improvement for clinical response; Yes for echocardiographic closure

## Discussion

This study provides real-world evidence from a tertiary neonatal intensive care unit, highlighting the clinical relevance of antenatal corticosteroids and gestational age in predicting treatment response and outcomes. These findings support early targeted screening and timely intervention in high-risk preterm infants.

In our cohort, the incidence of patent ductus arteriosus (PDA) was 2.8%, which is lower than rates commonly reported in the literature. This discrepancy may be explained by our strict inclusion criteria, excluding neonates without echocardiographic screening, those managed conservatively without pharmacological treatment (paracetamol or ibuprofen), and those with associated congenital heart disease. Furthermore, extremely preterm infants (<28 weeks of gestation), who are known to be at highest risk for PDA, were underrepresented due to the complexity of their management, which remains beyond the capacity of our setting.

Nearly 50% of mothers in our cohort presented with gestational toxemia, identified as the leading cause of medically induced prematurity. This finding is consistent with previous evidence suggesting that gestational toxemia is a significant risk factor for symptomatic PDA, primarily through its association with prematurity and impaired neonatal adaptation. A large retrospective study from the Korean Neonatal Network including 2961 preterm infants (22–29 weeks GA) reported a significant association between gestational toxemia and symptomatic PDA (P<0.001; OR=1.59 ; 95% CI : 1.24–2.04) [11].

The widespread use of antenatal corticosteroids has been associated with a reduction in PDA incidence, likely mediated by improved pulmonary maturation and reduced incidence of neonatal respiratory distress syndrome (NRDS), a well-established risk factor for PDA [11]. In our cohort, 95% of neonates presented with NRDS, highlighting the high baseline risk (Table 1). A Kuwaiti study by Hammoud et al. [12], including ventilated preterm infants <34 weeks GA (n=101), reported a markedly higher PDA incidence of 53.4%, underscoring the strong link between respiratory distress and PDA.

Only 50% of mothers in our study received antenatal corticosteroids, largely due to unanticipated preterm delivery. Among the 30 untreated mothers, 29 neonates developed symptomatic NRDS, emphasizing the protective role of antenatal steroids. This is supported by Shelton et al. [13], who demonstrated in a U.S. cohort (<27 weeks GA) that antenatal betamethasone significantly increased spontaneous ductal closure rates, reducing PDA incidence to 66% vs 84% in untreated infants.

Early targeted screening of high-risk neonates is increasingly advocated to enable pre-symptomatic treatment before day 3 of life [14]. The PDA-TOLERATE trial by Clyman et al. [15], conducted across 17 international centers, supports early intervention in infants <28 weeks GA. However, in our series, only 7 neonates underwent early screening, reflecting limited implementation of this strategy. Clinically, PDA suspicion was based on deterioration in respiratory and hemodynamic status, digestive intolerance, and characteristic murmurs, consistent with altered systemic perfusion due to ductal shunting (Table 2) [16].

Hemodynamic manifestations in our cohort included tachycardia (41%, n=25), widened pulse pressure (53.3%, n=32), fluid resuscitation (47%, n=28), and vasopressor use (30%, n=19). These findings align with a U.S. survey by Limrungsikul et al. [17], where hypotension, oliguria, systolic murmur, and cardiomegaly were the main indicators of hemodynamically significant PDA.

Altered cerebral perfusion associated with PDA contributes to the development of intraventricular hemorrhage (IVH) [6,18]. In our study, 22 neonates developed IVH. Similarly, Terrin et al. [8] demonstrated that hemodynamically significant PDA increased IVH risk by 3.184 in infants <28 weeks GA and 4.214 in those with birth weight <1000 g.

Echocardiography remains the gold standard for PDA diagnosis and therapeutic decision-making. Key parameters include ductal diameter, shunt magnitude, left atrium-to-aorta (LA/Ao) ratio, and systemic perfusion [6]. Most studies consider an LA/Ao ratio >1.15–1.7 or ductal diameter >1.5–2 mm as indicative of hemodynamic significance [19]. Kluckow and Evans [20] reported that a ductal diameter >1.5 mm predicts significant PDA with 83% sensitivity and 90% specificity. In our cohort, 58 neonates had a ductal diameter >1.5 mm, and 14 had a significant LA/Ao ratio (Table 3).

There is currently no universal consensus regarding optimal pharmacological management of PDA [5]. Ibuprofen remains the first-line treatment, with strong evidence supporting its efficacy. A Cochrane review (2018) demonstrated that intravenous ibuprofen significantly improves ductal closure rates compared to placebo [21], while other meta-analyses reported a reduced need for second-line therapy [19]. Cyclooxygenase inhibitors overall showed a failure risk ratio of 0.44 across 32 randomized trials [14].

Paracetamol has emerged as an alternative therapy since 2011, particularly in cases with contraindications to ibuprofen (e.g., sepsis, renal impairment, NEC) [22,23]. A systematic review updated in 2020 found comparable efficacy between oral paracetamol and intravenous ibuprofen, with fewer renal and gastrointestinal adverse effects in the paracetamol group [24].

In our study, ibuprofen was administered to 16 neonates (26.7%), while 44 (73.3%) received paracetamol. The overall ductal closure rate after first-line treatment was 83.3% (n=50), exceeding the 73% closure rate reported by Lago et al. [25].

Gestational age emerged as a key predictor of treatment success. Madan et al. [26] reported a significant correlation between gestational age and response to indomethacin, with closure rates of 43% (<23 weeks) versus 87% (27 weeks) (P=0.004; OR=1.51 per week). Similarly, in our study, closure rates were significantly higher in infants ≥28 weeks GA (P=0.003), with a 5.9-fold increase in success (Table 5).

Antenatal corticosteroids also improved treatment response. Chorne et al. [27] demonstrated an association between pulmonary maturity and successful PDA closure. In our cohort, corticosteroid exposure was a significant predictor of treatment success (P=0.011; OR=2.3) and echocardiographic closure (P=0.038; OR = 5.1; 95% CI: 0.96–26.9) (Tables 4-5). The wide confidence interval observed for antenatal lung maturation may be explained by the small sample size and the limited number of events, particularly in the group receiving lung maturation, leading to reduced precision of the estimate.

PDA-related complications in our study included bronchopulmonary dysplasia (BPD), IVH, and necrotizing enterocolitis (NEC). BPD occurred in 5% of cases and has been linked to prolonged PDA exposure [6,28]. A U.S. retrospective study (2020) confirmed this association in infants <29 weeks GA [28].

IVH occurred in 36% of our cohort, consistent with findings by Sellmer et al. [29], who reported an increased risk (OR=3.4) in infants <32 weeks GA, particularly when ductal diameter exceeded 1.5 mm. NEC was observed in 11% (n=7) of cases, likely due to reduced mesenteric perfusion associated with significant ductal shunting [5,10].

Mortality in our cohort was high (35%, n=21), with 8 deaths occurring in the first week of life.. This exceeds rates reported by Terrin et al. [8], where mortality was 10.4% vs 3.6% in PDA vs non-PDA groups (P=0.025). In our analysis, gestational age <28 weeks significantly increased mortality risk (P<0.001; OR≈5), while early clinical improvement reduced mortality by 3.7-fold (Table 6). Bussmann et al. [30] further demonstrated that prolonged PDA exposure is associated with increased mortality.

Our findings emphasize the importance of early echocardiographic screening in high-risk preterm infants to avoid delayed diagnosis of PDA. The strong association between antenatal corticosteroid exposure and improved outcomes highlights the need to optimize prenatal care. In addition, the high ductal closure rate (83.3%) supports the use of paracetamol as an effective alternative to ibuprofen, particularly in cases with contraindications. Finally, gestational age remains a key predictor of therapeutic success, supporting risk-adapted management strategies.

This study has several limitations. Its retrospective, single-center design may limit generalizability. The relatively small sample size reduces statistical power. Lack of systematic echocardiographic screening may have led to underestimation of PDA incidence. Additionally, potential inter-observer variability in echocardiographic measurements cannot be fully excluded. Treatment decisions were not standardized, introducing potential bias. Finally, the absence of long-term follow-up limits assessment of neurodevelopmental outcomes.

The findings of this study should be interpreted with caution due to its retrospective, single-center design, which may limit external validity. However, the study reflects real-world clinical practice in a tertiary neonatal intensive care unit and includes consecutive preterm infants, which enhances its applicability to similar high-risk settings. Caution is warranted when extrapolating these results to different populations, particularly extremely preterm infants or centers with different management protocols.

Prospective multicenter studies with larger sample sizes are needed to confirm these findings. Further randomized trials comparing paracetamol and ibuprofen are required to define optimal treatment strategies. Future research should also focus on standardized screening protocols, early treatment strategies, and long-term neurodevelopmental outcomes in preterm infants with PDA.

## Conclusion

Patent ductus arteriosus remains a significant cause of morbidity and mortality in preterm infants. Early identification, antenatal corticosteroid exposure, and timely pharmacological treatment are key to improving outcomes. Implementation of systematic echocardiographic screening in high-risk neonates may facilitate earlier diagnosis and management.

## Declarations

### Competing Interests

The authors declare that they have no competing interests.

## Funding

This research did not receive any specific grant from funding agencies in the public, commercial, or not-for-profit sectors.

## Data Availability

The minimal dataset underlying the findings of this study is available from the corresponding author, Hayet Ben Hamida, MD, upon reasonable request. Access to the data will be provided in compliance with patient confidentiality and ethical regulations.

## Acknowledgments

The authors thank the medical and nursing staff of the Neonatal Intensive Care Unit of the Teaching Hospital of Monastir for their dedication to patient care. We also acknowledge the Department of Cardiology B for their contribution to echocardiographic assessment, and the hospital administration for supporting clinical research activities.

## Authors’ Contributions

Hayet Ben Hamida : Conceptualization, Methodology, Data curation, Formal analysis, Investigation, Writing – original draft.

Maroua El Ouaer: Data curation, Investigation, Writing – review & editing.

Sioir Abdelmoula : Investigation, Data curation, Writing – review & editing.

Maha El Ghali : Investigation, Data curation, Writing – review & editing.

Manel Bizid : Data curation, Software, Writing – review & editing.

Ikram Chamtouri : Investigation, Methodology, Writing – review & editing.

Kamel Monastiri : Conceptualization, Methodology, Supervision, Writing – review & editing.

All authors have read and approved the final version of the manuscript.

## Notes

### Competing Interest Statement

The authors have declared no competing interest.

### Clinical Trial

?Not applicable?

### Funding Statement

The author(s) received no specific funding for this work.

### Author Declarations

The study was approved by the Institutional Review Board of the Faculty of Medicine of Monastir and was conducted in accordance with ethical standards for research involving human participants. All patient data were handled anonymously and in compliance with confidentiality regulations.

